# Dysmetabolic circulating tumor cells are prognostic in metastatic breast cancer

**DOI:** 10.1101/19012211

**Authors:** Giulia Brisotto, Eva Biscontin, Elisabetta Rossi, Michela Bulfoni, Aigars Piruska, Simon Spazzapan, Cristina Poggiana, Riccardo Vidotto, Agostino Steffan, Alfonso Colombatti, Wilhelm T. S. Huck, Daniela Cesselli, Rita Zamarchi, Matteo Turetta, Fabio Del Ben

**Author notes:** These authors contributed equally to this work (Giulia Brisotto, Eva Biscontin). These authors jointly supervised this work (Matteo Turetta, Fabio Del Ben).

## Abstract

The prognostic value of the circulating tumor cells (CTCs), defined as EpCAM+, Cytokeratin (8, 18, 19)+ and CD45-nucleated cells, has been provided in metastatic breast cancer (mBC), with Level I of evidence. However, CTCs belong to a heterogeneous pool of rare cells, and there isn’t consensus on an univocal definition of CTCs. Here, we present a definition of metabolically altered CTCs (MBA-CTC) as CD45-negative cells with an increased extracellular acidification rate (iECAR), supported by the presence of iECAR among the hallmarks of cancer. We tested the prognostic value of MBA-CTC present in mBC patients before starting a new systemic therapy (T0) and 3-4 weeks after (T1). Samples were analyzed in parallel with CellSearch platform (CS). Standard RECIST criteria were used to determine patients’ responses to treatment.

In our cohort of n=31 mBC patients, a level of MBA-CTCs above the cut-off was associated with: i) a shorter median PFS both pre-therapy (123 days vs 306; p<0.0001) and during therapy (139 vs 266 days; p= 0.0009); ii) a worse OS both pre-therapy (p=0.0003, 82% survival vs 20%) and during therapy (p=0.0301, 67% survival vs 38%); iii) good agreement with therapy response (kappa=0.685). Both the trend of MBA-CTCs over time and the combined results of the two assays (MBA and CS) enabled more accurate stratification. MBA and CS results showed fair (K=0.33) and poor (K=0.077) agreement at T0 and T1, respectively. This fact and the increased accuracy in combining results suggest that the assays detect different CTC subsets. In conclusion, MBA-CTCs does provide prognostic information at least equivalent to CS, and are even more informative when analyzed over time or combined with CS-CTCs.

## INTRODUCTION

Breast cancer is the most common cancer among women worldwide, with an estimated 2 million of newly diagnosed cases each year *(1)*. Despite the great advances in the early diagnosis and treatment, metastatic spread still represents the leading cause of breast cancer-related death *(1)*. The availability of effective biomarkers able to stratify the patient population which will most likely benefit from a specific therapy, monitor the treatment response, quantify the minimal residual disease, and promptly detect the appearance of therapy resistance, still represents a clinical need *(2)*.

Circulating tumor cells (CTCs) are cancer cells spread from primary tumor or metastatic sites and found in body fluids. Their analysis has gained attention as one of the most promising approaches to face the aforesaid needs in the clinical management of cancer, as CTCs constitute tumor material that can be accessed with minimal invasiveness, providing real-time information on the evolving biology of cancer *(3–7)*. There is evidence that CTCs can contribute to form distant metastasis *(8)*, and several studies demonstrated that the number of CTCs independently correlates with progression of disease in metastatic breast cancer (mBC) patients, outperforming both serum biomarkers *(9, 10)* and radiologic imaging techniques *(11)*.

The only currently FDA-approved technology for CTC enumeration in mBC is the CellSearch platform (Menarini) that detects CTCs relying on the expression of the epithelial marker EpCAM on their surface. This technology has widely demonstrated the prognostic value of both the number *(12–15)* and number variation *(11, 16)* of CTCs, in patients with early and metastatic BC.

CellSearch and other similar technologies, though, are limited to the detection of CTCs expressing the epithelial marker EpCAM. However, the invasion of tumor cells from peripheral blood is a complex process, during which tumor cells can undergo epithelial-to-mesenchymal transition (EMT) that involves the downregulation of epithelial markers, such as EpCAM, cytokeratins and E-cadherin, and the upregulation of mesenchymal markers, such as Vimentin and N-cadherin. Several groups have shown the existence of non-epithelial CTCs and linked these subtypes to disease progression or other clinical-pathological features *(17–21)*. A plethora of new technologies emerged to overcome the limitations of EpCAM-based approaches, focusing on the challenge of detecting non-epithelial CTCs, mainly exploiting mesenchymal markers or physical features of cancer cells and other cancer-specific traits. Notably, few groups have specifically addressed altered metabolism of cancer cells *(22–27)* – recognized as a “hallmark of cancer”*(28)* – as a promising approach, also in view of the introduction of specific metabolic agent combined with chemo- or immunotherapies in several clinical trials *(29–31)*. One of the most described metabolic alterations of cancer cells consists of a markedly increased uptake of glucose in comparison with normal cells. First described by Otto Warburg in the 1920s, this cancer feature has been successfully exploited in the clinic by introducing a positron emission tomography (PET)-based imaging of the uptake of a radioactive glucose analog, which is commonly employed to stage cancer and assess response to therapy *(32)*. Several groups reported the exploitation of metabolic alterations to identify CTCs in the peripheral blood of metastatic cancer patients, independently from EpCAM expression and immunostaining techniques in general *(22–27)*. An mRNAseq study on CTCs from prostate cancer patients described a category of CTCs with an upregulation of metabolic transcripts as a potential biomarkers for cancer metastasis *(24)*. Tang et al *(22)* evaluated the presence of metabolically active CTCs in pleural effusion and in the peripheral blood of a limited number of non-small cell lung cancer patients by using the fluorescent glucose analogue 2-NBDG. This finding was also strengthened by our group in a preliminary study to assess the mutational status of hypermetabolic CTCs enriched by their elevated 2-NBDG uptake *(23)*. Other microfluidic platforms have been described for the screening of single-cell uptake of glucose or the release of by-product of metabolism such as lactate *(33)* or ROS *(34)*.

In this study we focused on pH dysregulation *(35–37)*: in cancer, an augmented extracellular acidification is the consequence of both the increased lactate extrusion (in the context of the Warburg phenomenon) and of the sodium-proton antiporter NHE1 hyperactivity. pH dysregulation, an early phenomenon in carcinogenesis, is important for cancer cells to gain selective advantages and a more metastatic phenotype, and the acidic pH of the tumor milieu plays a recognized role in cancer invasion, metastasis, immunosuppression and therapeutic resistance *(35)*. Therefore, an abnormal metabolism constitutes an excellent candidate as a functional marker: since it represents an independent feature with respect to epithelial and mesenchymal markers, in principle, it could be leveraged for CTC detection without the need to identify specific phenotypic profile.

Droplet-based microfluidic approaches are typically used for the encapsulation of cells, as they allow to maintain cell growth and proliferation properties and to screen for secreted molecules, as they are retained inside the droplet *(38, 39)*. We previously demonstrated that the differential release rate of protons between cancer cell lines and normal white blood cells (WBCs) can be detected in-drop exploiting a pH-sensitive dye, which allows to assess their increased extracellular acidification rate (ECAR) by measuring their acidification dynamic *(25)*. As a proof of concept, we demonstrated the ability of the pH value to discriminate cancer cell lines from WBCs, and to detect CD45-negative cells with increased ECAR in patients with metastatic cancer of different types *(25)*.

In this paper, we refer to this CTC detection method as “metabolism-based assay” (MBA). Here, we determined the prognostic role of MBA-CTCs in mBC and compared their performance with epithelial CTCs, as detected by the gold-standard CellSearch assay (CS-CTCs).

## RESULTS

### Patients’ characteristics

Between September 2016 and December 2017, 31 consecutive mBC patients were recruited at the Aviano National Cancer Institute, regardless the number and type of previous lines of treatment. CTCs were enumerated using both the metabolic-based assay (MBA) and CellSearch assay (CS) prior to the beginning of a new line of treatment (T0) and after a median of 3 weeks following the first cycle of therapy (T1). The CONSORT diagram of evaluable patients for each method is shown in fig. S1. Patient demographics and tumor characteristics at the time of enrollment are summarized in Table 1. The patients’ median age was 56. Primary tumor receptor status for ER and/or PR (detected by IHC) and HER2 (evaluated by IHC or FISH) was positive in 21 (68%) and 3 (10%) out of 31 patients, respectively, while 8 (26%) out of 31 cases were triple-negative (ER-, PR- and HER2-negative).

**Table 1.** Patient and tumor characteristics

|  | N | % |
| --- | --- | --- |
| **All patients** | 31 | 100.0 |
| **Progressed** | 25 | 80.6 |
| **Alive** | 18 | 58.0 |
| **Age at baseline (years)** |  |  |
| Median | 56 |  |
| Range | 39-78 |  |
| **ER- and PR-receptor status** |  |  |
| ER+ or PR+ | 21 | 68.0 |
| ER- and PR- | 10 | 32.0 |
| **Her2/neu status** |  |  |
| Positive | 3 | 10.0 |
| Negative | 28 | 90.0 |
| **Triple negative** | 8 | 26.0 |
| **Sites of metastasis** |  |  |
| Visceral | 21 | 68.0 |
| Non-visceral | 10 | 32.0 |
| Bone | 23 | 74.0 |
| Lung | 14 | 45.0 |
| Brain | 2 | 6.0 |
| Liver | 14 | 45.0 |
| Nodes | 19 | 61.0 |
| **Number of metastasis** |  |  |
| 1 | 9 | 29.0 |
| 2 or more | 22 | 71.0 |
| **Therapy** |  |  |
| First line | 11 | 35.5 |
| Second line or subsequent | 20 | 65 |
| **Type of therapy** |  |  |
| Chemotherapy alone | 20 | 64.5 |
| Chemotherapy and targeted therapy | 8 | 25.8 |
| Hormone-therapy | 2 | 6.5 |
| Placebo | 1 | 3.2 |

Among the 31 patients recruited, 11 (35.5%) were starting their first line of therapy for metastatic disease; 28 (90.0%) received chemotherapy (alone or in combination with other treatments), 2 (6.5%) received hormone-therapy and 1 (3.2%) patient did not receive any treatment (control group of the A-BRAVE clinical trial, NCT02926196).

All patients had a minimal follow-up time of 12 months (median 18 months), and the first follow-up imaging study was performed on average 5.3 ± 2 months after the T0 blood sample collection. First imaging re-evaluation after CTC enumeration documented a partial response/stable disease in 13 (42%) out of 31 patients; after the whole period of follow-up, instead, disease progression was observed in 25 (80.6%) out of 31 cases, and 13 (42.0%) patients deceased. Blood samples from 26 healthy donor (HD) volunteers were analyzed with MBA as negative controls.

### MBA description

The overall strategy adopted to detect cancer cells in a background of WBCs, by measuring the extracellular acidification rate (ECAR) *(25)*, is outlined in Fig. 1 and fig. S2. Briefly, single cells were encapsulated into 15 pL droplets together with a fluorescent pH-sensitive dye (SNARF-5F) (Fig. 1A). During in-drop incubation, because of the cellular metabolic activity, each cell extruded a certain quantity of H^+^ altering the pH of the droplet (Fig. 1B), which was then quantified by optical readout (Fig. 1C and fig. S2). Beside pH, the system allowed to determine EpCAM and CD45 expression, as well as to record images of analyzed droplets to confirm the presence of a single cell within the droplet (fig. S2).

**Fig. 1.**
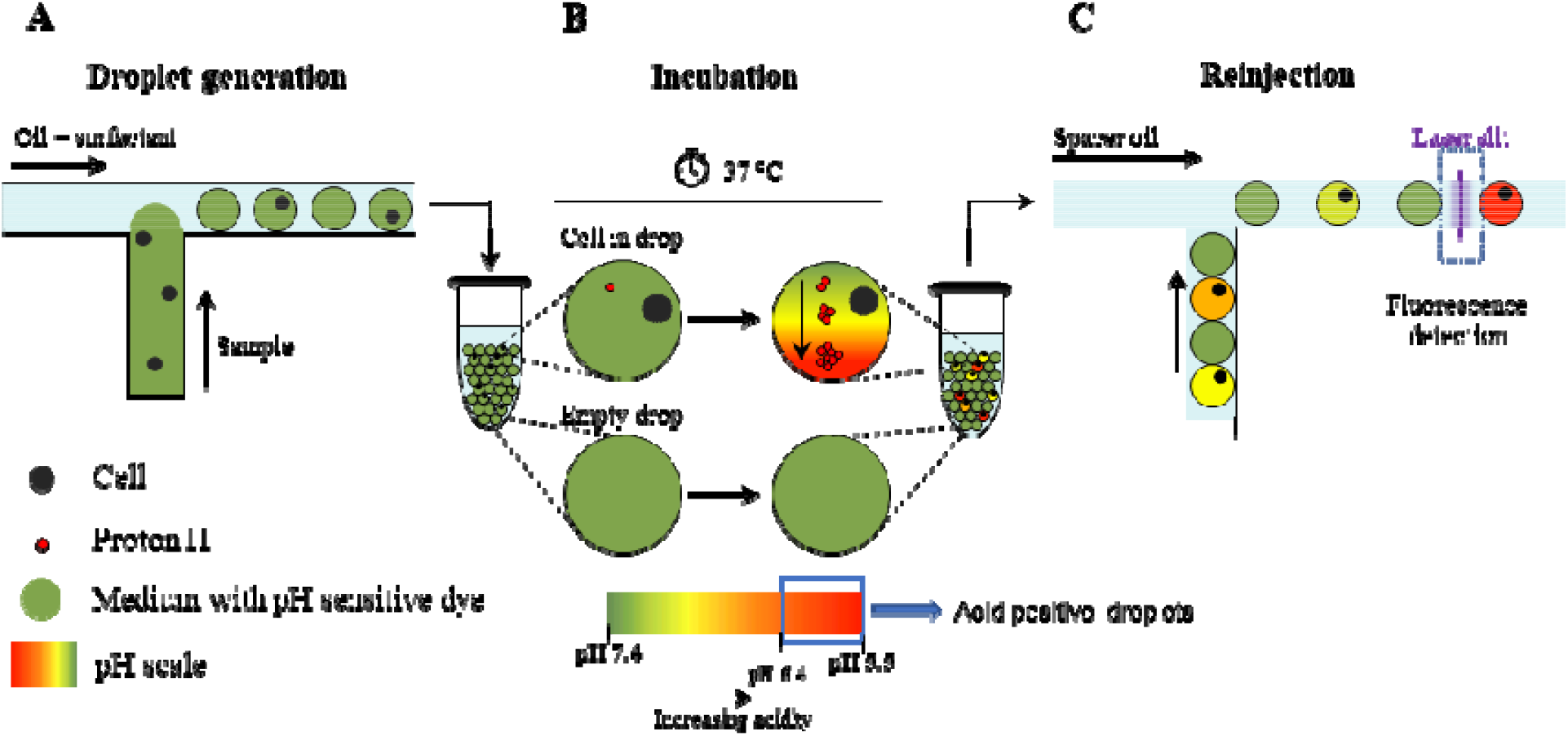
Schematic overview of the droplet microfluidic single-cell MBA for determination of the extracellular pH. **(A)** Single-cells are encapsulated into picoliter droplets together with an extracellular pH-sensitive dye (SNARF-5F); **(B)** During in-drop incubation, single cells secrete a certain quantity of protons (H+)which leads to a decrease of the pH value (i.e.: acidification of the droplet content); **(C)** Droplets are reinjected into a second microfluidic device and screened for fluorescence by the optical setup.

To determine a pH cut-off value for detecting breast cancer cells in mBC patients, both a basal-like, triple-negative (MDA-MB-231) and a luminal, hormone receptor positive (MCF-7) breast cancer cell lines, which are considered, respectively, representative of EpCAM-low and EpCAM-high *(40)* cell lines, were tested. By comparing breast cancer cells with WBCs, the cut-off value of pH<6.4 gra ted detection of both breast cancer cell lines with a negligible number of WBC contaminants (0.045%, percentage which is further reduced in the patient analysis by CD45-labeling) (fig. S3). Then, in subsequent analyses, cells that were negative for CD45 expression and within a droplet with pH<6.4 were considered as MBA-CTCs.

In *ex-vivo* mBC blood samples, CTC count has been used to categorize patients as negative or positive for the presence of CTCs, if their count was 0 or ≥ 1, respectively. Moreover, a cut-off of 6 CTCs per 7.5 ml of blood was determined, adopting the same strategy that was used to set at 5 cells the prognostic cut-off value for the CS assay *(10, 15)* (see Materials and Methods and fig. S4 for details); accordingly, patients were stratified as MBA-CTC <6 and MBA-CTC ≥6.

### Baseline CTC enumeration

Using the MBA assay, at T0, CTCs were detected in 12 out of 27 patients (44.4%), with 10 patients (37.0 %) being above the cut-off value of 6 cells. Overall, the average CTC count was 218±1022 (median 0, range 0-5319) (Table 2 and table S1).

**Table 2.**
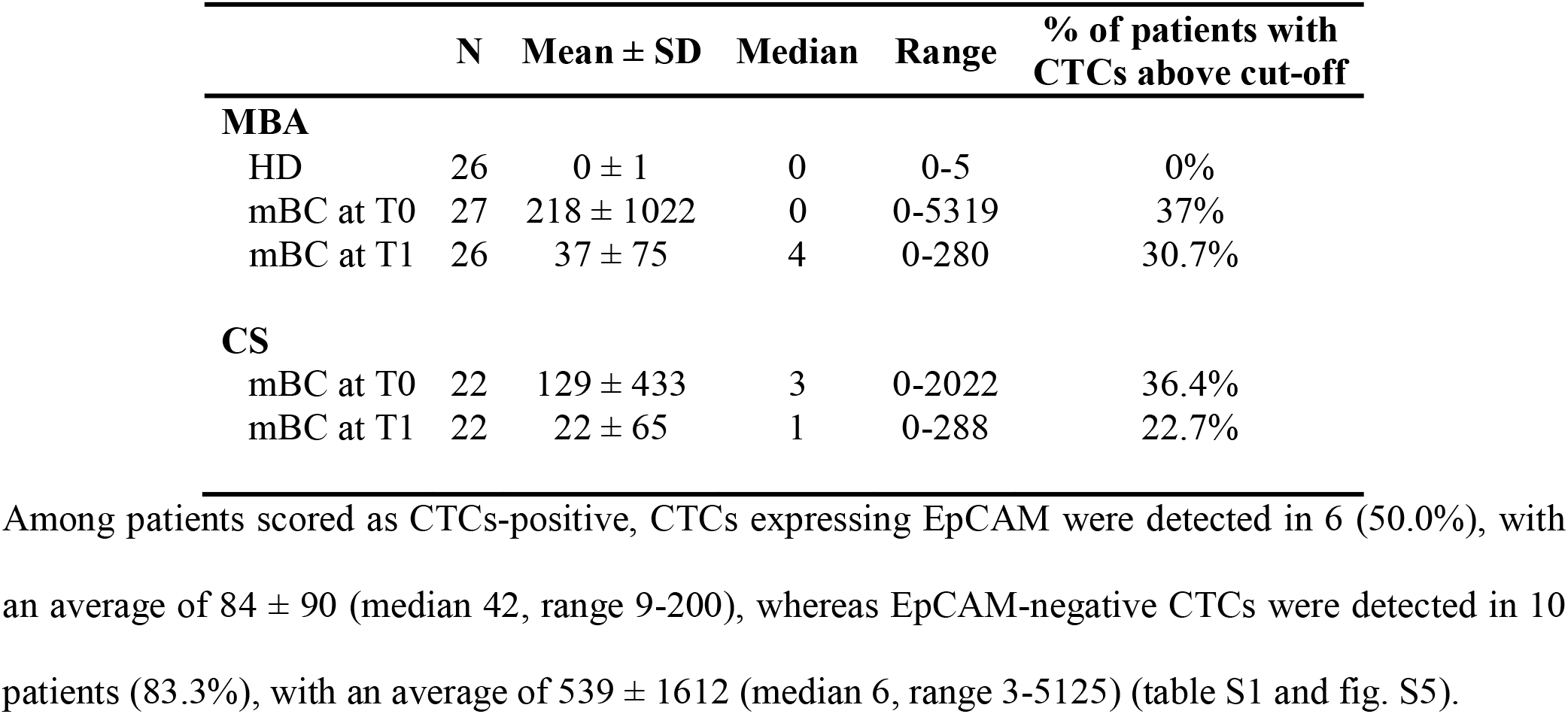
Prevalence of CTCs at baseline (T0) and follow-up (T1) as detected by the metabolism-based assay (MBA) and CellSearch (CS), according to the cut-off of 6 and 5 CTCs, respectively. (HD = healthy donor volunteers; mBC = metastatic breast cancer patients).

In the cohort of HD volunteers, used as negative controls in this study, events with a metabolic signal comparable to CTCs could be detected in 2 (7.6%) out of 26, showing 4 and 5 cells, respectively (Table 2 and table S2). Please note that these values are obtained by normalizing analysed sample volume to 7.5 mL for easier comparison with CellSearch, as described in the methods section. These two values correspond to 1 false positive event detected in each of these two HD in 1.9 and 1.5 mL of sample analyzed, respectively. Overall, HD cohort showed 2 false-positive events in more than 50 millions of droplets analysed. Notably, none of the HD showed more than 6 cells and the number of CTCs was significantly higher in mBC patients than in HDs (p=0.001, Mann-Whitney U-test) (fig. S5).

By analyzing the same mBC population by CS, we found that 17 (77.3%) out of 22 evaluable patients had at least 1 CTC, while 8 (36.0%) patients had a CTC number above the prognostic cut-off of 5 cells (as established in previous clinical studies) *(10, 15)*. Overall, the average number of CTC was 129±433 (median 3, range 0-2022).

Moreover, we also determined the level of apoptotic CTCs by CS platform, as previously reported *(41)*, by detecting in CTCs the expression of M30, an epitope of cytokeratin 18 revealed during early phase of apoptosis. Among the CTC-positive patients, 3 (17.6 %) had at least 1 M30-positive CTC with an average count of 9 ± 14 (median 1, range 1-25) (table S1).

### Changes of CTC levels after starting treatment

By the MBA, 15 (57.7%) out of 26 patients presented at least 1 CTC, while 8 (30.7%) had more than 6 cells. With respect to T0, the overall average number of CTC decreased from 218±1022 to 37±75 (median 4, range 0-280) (Table 2 and table S1). EpCAM-positive cells were found in 7 (46.6%) of MBA-CTC positive patients, with an average of 48±65 (median 18, range 4-160) cells, whereas patients presenting EpCAM-negative CTC were 12 (80%) out of 15, with an average of 52±70 (median 14, range 3-225) cells (table S1 and fig. S5).

Comparing CTC number at T0 and T1 for each patient, CTC level decreased in 10 cases and increased in 6 (fig. S6), while 7 patients were negative at both time-points. The CTC concentration at T0 did not statistically differ respect to T1 (p=0.2465, Wilcoxon test).

By CS analysis, at T1, 11 (50%) out of 22 evaluable patients had at least 1 detectable CTC, whereas, 5 (23%) out 22 patients showed ≥5 CTCs. The average CTC number decreased from 129±433 to 22±65 (median 1, range 0-288) (Table 2 and S1).

Moreover, among the CTC-positive patients, apoptotic CTCs (M30-positive) were detected in 2 (18,2%) out of 11 sample, accounting for 1 and 7 cells, respectively. Overall, the number of M30-positive cells at both time-points was so low that it did not allow reliable data analysis.

Among the 17 patients that had CS paired samples at T0 and T1, 2 (11,8%) had an increase and 10 (58,8%) a decrease in CTC levels, while 5 (29,4%) cases showed unchanged CTC value (fig. S6). Unlike MBA results, CS-CTC levels significantly differed between T0 and T1 (p=0.0146, Wilcoxon test).

### Comparison of CTC levels using MBA and CS

A total of 22 patients were analyzed in parallel with both the MBA and the CS at T0 and 21 patients at T1. The total number of patients with a CTC level < or ≥ the cut-off for each technique at each time-point is reported in Table 3. Using the respective prognostic cut-offs (MBA: ≥6 CTCs; CS: ≥5 CTCs), the overall positive concordance was 68.2% at T0 and 61.9% at T1; the correlation between matched samples was weak at T0 and absent at T1 (T0: Spearman r =0.39; T1: Spearman r = 0.04) (Fig. 2A and B); this finding combined with a low k-Cohen coefficient (Table 3) indicated low agreement between the two methods.

**Table 3.**
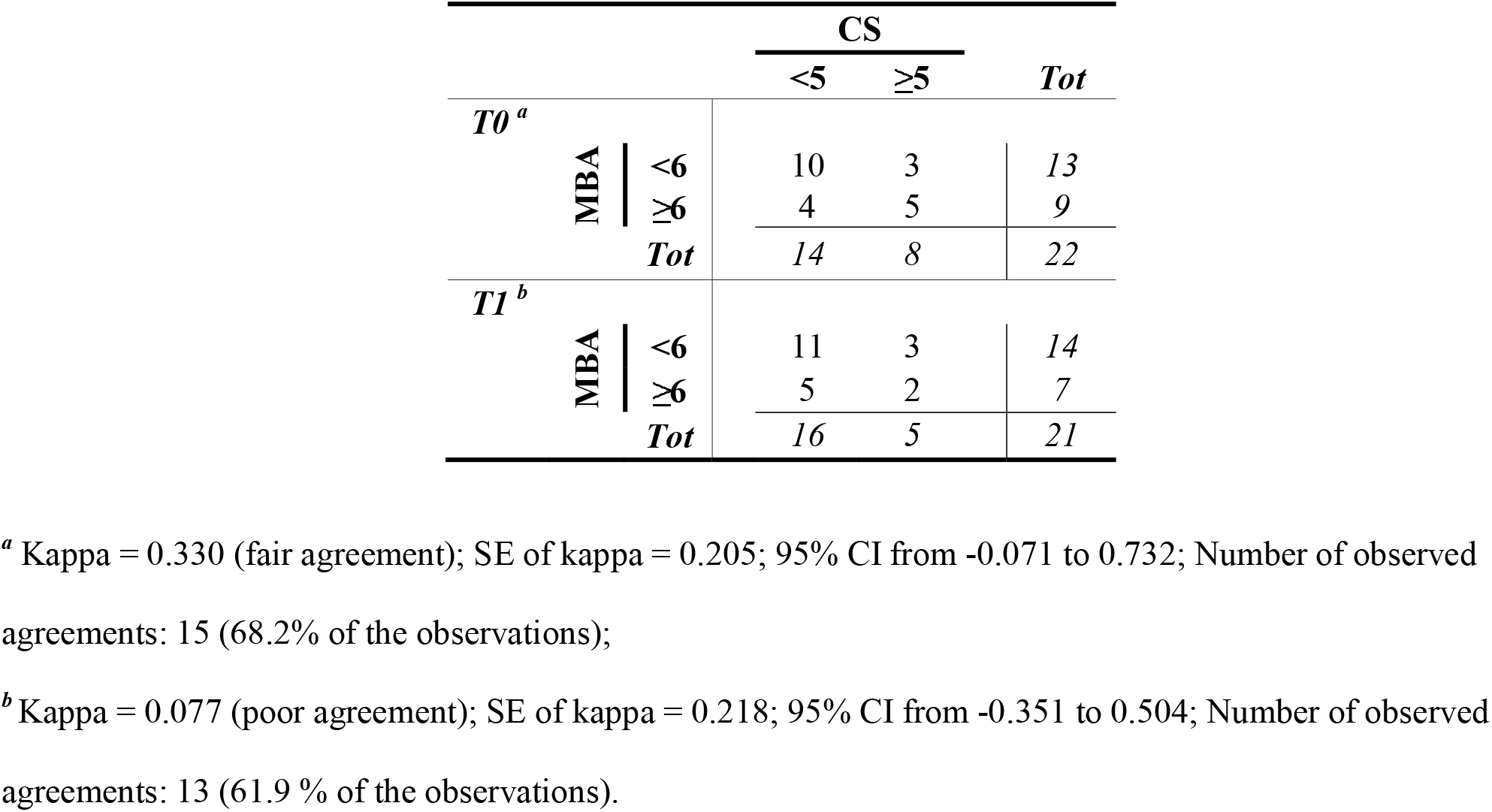
Concordance of CTC status between the metabolism-based assay (MBA) and CellSearch (CS) method at baseline (T0) and follow-up (T1), using respective cut-offs.

**Fig. 2.**
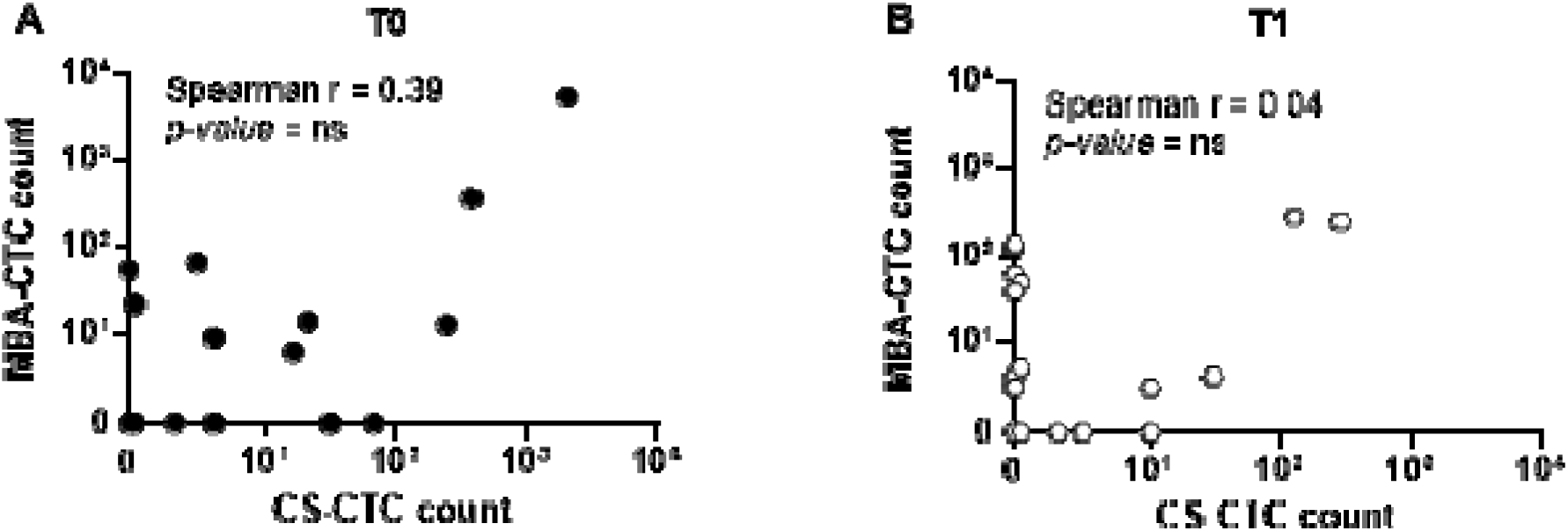
Direct comparison of CTC enumeration between the MBA and the CellSearch method. **(A)** Correlation of CTC count as detected by the MBA and the CS method at baseline (T0) and **(B)** follow-up (T1).Only complete pairs of data including results from both MBA and the CS method were plotted. Overall, 22 and 21 cases had matched blood sample at T0 and T1, respectively.

### Concordance between CTC status and radiologic imaging

The agreement between the number of CTCs assessed at T0 and T1 and the presence of stable disease/partial response (SD/PR) or progressive disease (PD) at the first follow-up imaging study was tested. For MBA-CTCs, a good agreement (kappa= 0.685) at T0 and a fair agreement (kappa=0.306) at T1 were observed. CS showed a moderate agreement both at T0 (kappa=0.441) and at T1 (kappa=0.431). In the best case (MBA-CTC at T0), 84.6% of therapy responses was correctly predicted by MBA. Results are shown in Fig. 3 and table S3.

**Fig. 3.**
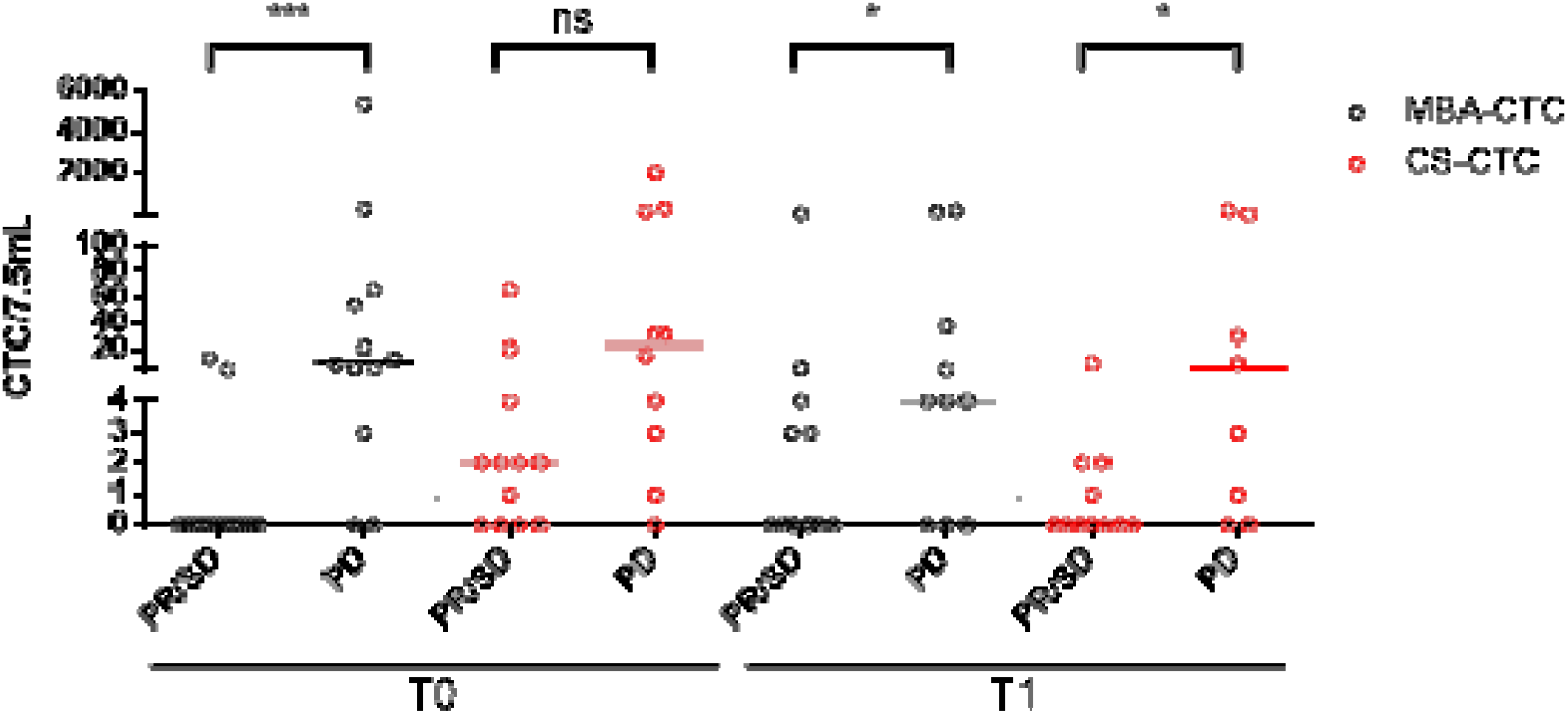
Comparison between CTC level as detected by the metabolic-based assay (MBA) and CellSearch(CS) and imaging. This plot shows the CTC count for individual patients at baseline (T0) and follow-up (T1), compared with their response at the first follow-up imaging. (PR/SD = Partial Response/Stable Disease; PD =Progressive Disease. Statistical significance was calculated by Mann-Whitney test (*** p<0.001, * 0.01<p<0.05, ns p>0.05).

### Survival analysis

As shown in Fig. 4, progression-free (PFS) and overall survival (OS) were predicted at T0 and T1 for both methods, when patients were stratified according to the respective prognostic cut-off values. Patients analyzed by MBA and stratified according to CTC number ≥6 or <6, showed a significantly different median PFS at both T0 (123 vs 306 days, p<0.0001) and T1 (139 vs 266 days, p= 0.009) (Fig. 4A and B). Similarly, MBA-CTC level ≥6 or <6 was able to predict OS at both T0 and T1 (T0: 243 vs >600 days, p=0.0003; T1: 209 vs >600 days, p=0.03) (Fig. 4C and D). Conversely, the CS results were only partially overlapping with MBA data. Indeed, the CS-CTC number ≥5 or <5 predicted PFS (T0:131 vs 262 days, p=0.0146; T1: 119 vs 260 days, p= 0.0152) (Fig. 4A and B) at both the examined time-points, and OS at T0 (227 vs >600 days, p=0.019) but not at T1 (191 vs >600 days, p=0.0636) (Fig. 4C and D). Notably, an exceptionally high number of CTCs characterized two patients with an extremely short survival, at both time-points with both techniques.

**Fig. 4.**
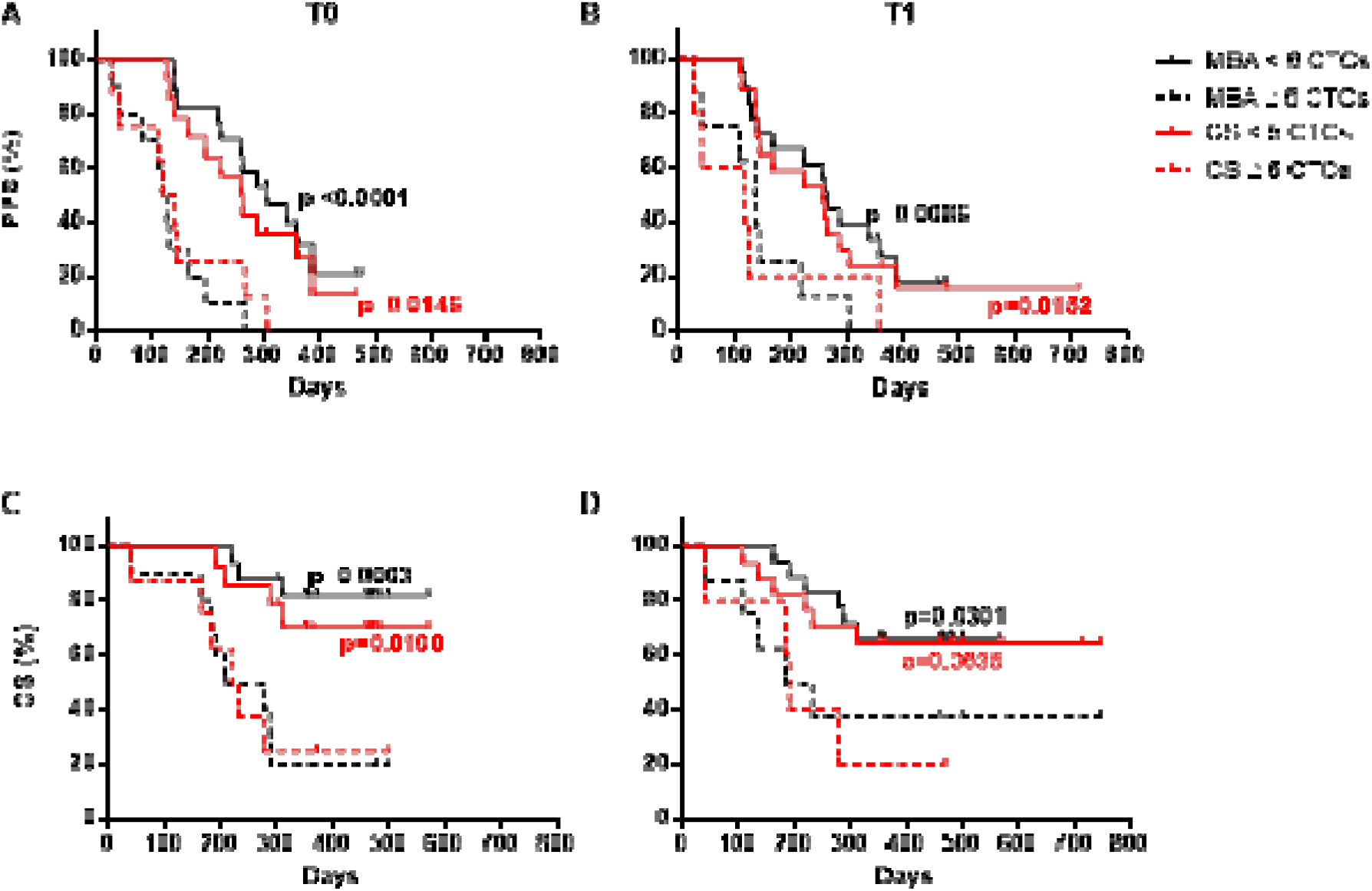
Progression-free survival (PFS) and overall survival (OS) of mBC patients. **(A)** Kaplan-Meier plots estimating PFS at baseline (T0) and **(B)** first follow-up (T1) by the metabolic-based assay (MBA) and the CellSearch (CS) method. **(C)** Kaplan-Meier plots estimating OS at baseline (T0) and **(B)** first follow-up (T1) by the MBA and the CS method. For these analyses, mBC patients were stratified using a cut-off value of 6 CTCs for the MBA and 5 CTCs for the CS.

## DISCUSSION

Studies of other groups involving CTC detected with a metabolism-based approach did not correlate results with clinical outcome. In the present pilot study, we determined that elevated MBA-CTCs are linked to shorter overall and progression-free survival. Additional information was obtained considering CTC levels over time.

One of the limitations of this study is the relatively small cohort involved, though our cohort reflects general population in terms of cancer subtype prevalence (luminal, HER2-positive, triple-negative), metastatic sites and lines of therapy. This is further confirmed by the results obtained in this study with CS, which are aligned with larger clinical trials in metastatic breast, colon and prostate cancers *(10, 11, 15, 16, 42, 43)*. Despite relatively small cohort results display unequivocal statistical significance. Thus, we feel confident enough stating that the presented results offer robust evidence that MBA-CTCs are prognostically informative across different subtypes of mBC patients. Another limitation is the relatively small volume of sample analyzed (2.5mL vs usual 7.5mL), which could have lead to a slight underestimation of MBA-CTC. The limit of detection is bigger if a smaller volume is analyzed. Specifically, the limit of detection was 1 MBA-CTC in 2.5 mL, corresponding to 3 MBA-CTC in 7.5 mL. In other words, the presence of <3 MBA-CTC in 7.5 mL might have led to a result 0 MBA-CTC in 2.5 mL, thus falsely underestimating the actual number of CTC. However, this did not affect survival curves because 3 MBA-CTC is smaller than the cut-off used to stratify patients (MBA-CTC < 6). Interestingly, the agreement between MBA and CS was relatively weak, while both methods independently yielded similar prognostic significance. This suggests that the two techniques detect different CTC subgroups with complementary information, possibly reflecting different aspects of disease evolution. By combining information of the two assays, additional information could be gained. This interpretation is strengthened by the fact that the prevalence of single-positive patients (either MBA or CS above cut-off) is roughly 50% higher than the prevalence obtained considering single MBA or CS testing. A possible explanation is the fact that metabolism is altered also in non-epithelial, which are mostly EpCAM-negative and thus not detected by CellSearch. At the same time, our pH threshold was relatively stringent in order to maximize purity, since it is set in order to obtain absence of dysmetabolic cells in healthy controls or absence of white blood cells contaminants in other words. This could lead to “missing” of some EpCAM-positive cells with increased ECAR with respect to normal WBC, but not enough increased to meet pH threshold.

It should be now clear that referring to CTCs without further definition might be confusing, as it is a rather blurred group of cells of different nature and maybe distinct clinical implications. More studies are needed to clearly understand if different types of cells could be linked to specific clinical and pathological features or therapy susceptibilities.

Hypothesizing a predictive role of MBA-CTC, a growing body of evidence suggests that the acidic extracellular environment of tumor tissues confers chemoresistance: indeed, a majority of chemotherapeutics (e.g.: doxorubicin) behaves like weak bases which are quickly protonated in low pH environments, leading to a reduced cellular uptake and treatment efficacy *(44)*. Thus, restoring a physiologic extracellular pH is emerging as a new therapeutic strategy to increase tumor chemosensitivity to traditional chemotherapeutics. Several pH regulator inhibitors have already been evaluated in several clinical trials *(30, 45)* and, in this contest, MBA-CTC might represent a fitting approach to monitor the benefit of this new therapeutic strategy.

## CONCLUSION

CTC identified by their aberrant metabolism are at least as tightly linked to survival and disease progression as epithelial CTCs, either when measured before or after the therapy. Overall, this pilot study can be used as a foundation to design larger clinical studies to confirm and validate MBA-CTCs as a prognostic biomarker.

## MATERIALS AND METHODS

### Study design

A pilot study trial was conducted adopting both the metabolism-based assay (MBA) and the CellSearch system for CTC enumeration. The study was conducted at the IRCCS-CRO Aviano-National Cancer Institute and approved by our Institutional Review Board. Informed and written consent was obtained from all patients and healthy donors before their enrolment, and their clinico-pathological information was recorded. A total of 31 patients with progressive and measurable mBC, at the start of a new systemic therapy, without limits to number and kind of previous therapies (hormone therapy, chemotherapy, targeted therapy) were included. All patients had an Eastern Cooperative Oncology Group performance status (ECOG PS) score ≤ 1.

Before starting a new therapy, patients underwent a baseline (T0) blood drawn for CTC evaluation and routine clinical tests. Another blood sample was collected 3-4 weeks after the beginning of the therapy (follow-up, T1). Among the 31 enrolled patients, 27 and 26 blood samples were evaluated for the presence of CTC with the MBA at T0 and T1, respectively, whereas CTC count was evaluable with the gold standard CellSearch for 22 patients at both time points. Details about reasons for exclusion (e.g.: insufficient or clotted blood sample, organizational and/or technical failure) are summarized in fig. S3. Clinical re-evaluation of the disease status was conducted depending on the type and schedule of the therapy; Standard Response Evaluation Criteria In Solid Tumors (RECIST) criteria were used to determine patients’ responses to treatment.

### Microfluidic platform

#### Device fabrication

In this work, a microfluidic device was employed for droplet generation, and a second one for droplet reinjection in order to screen their fluorescence for pH determination and antibody staining. Each device was made of PDMS bonded to a glass surface, as previously reported^27^.

#### Optical setup

The optical setup for measuring droplet fluorescence consisted of an Olympus IX70 inverted fluorescence microscope. A 25mW, 405 nm laser beam was focused with a cylindrical lens crossing orthogonally the microfluidic channel. Fluorescence signal emitted from droplets was captured by a 40x objective (Olympus LUCPlanFLN, 40x/0.60), split with dichroic filter and detected through bandpass filters (579/34; 630/38 e 494/20, 435/20) by Photo Multiplier Tubes (PMTs) (H957-15, Hamamatsu). Signal went through a transimpedance amplifier with 1V/uA gain and detected by the acquisition system (National Instruments cRIO-9024, analog input module NI9223) with a 10 μsec scan rate.

#### Droplet generation and encapsulation of cells

The emulsification device was used to generate droplets. This device has a flow-focusing junction, where the aqueous phase (cell suspension) and carrier oil stream meet, and droplet generation occurs (forming a water-in-oil emulsion). The resulting emulsion flowed off-chip through PTFE tubing (Fisher Bioblock) connected to the outlet of the device and collected into 1.5ml vial (Eppendorf) placed on an ice-cold rack (IsoTherm System, Eppendorf). The vial containing droplets was incubated for the desired incubation time at 37°C. After incubation, droplets were immediately cooled by replacing the vial on the ice-cold rack and reinjected into the required microfluidic device for subsequent analysis.

#### Droplet fluorescence screening, data acquisition and control system

Droplets were reinjected into the emulsion device through PTFE tubing (Fisher Bioblock). The data acquisition system has a 10 μsec scan rate. The signal output voltage of each droplet was recorded and processed in real time by a Field Programmable Gate Array (FPGA) card. Droplets with a size within defined range and gated fluorescence were imaged by synchronous triggering of an LED and a CMOS camera, and (optionally) sorted with a dielectrophoretic pulse. Data are transferred from the FPGA to the real-time controller providing a record of images and raw tracks for each run. A custom LabVIEW software allows the operator to access the picture gallery, evaluate the pictures of droplets to discard droplets that are empty or containing particles not compatible with the morphology of intact cells (debris).

### pH-assay for extracellular acidification measurements

The pH-sensitive fluorescent dye SNARF-5F (Invitrogen) was used to measure the pH of each droplet. SNARF-5F respond to pH variation undergoing a wavelength shift in the emission spectra. Such pH-dependent shifts allow the ratio of the fluorescence intensities from the dye at two emission wavelengths (580nm and 630nm) to be used for quantitative determination of pH. For each droplet the ratio of emitted fluorescence intensities at 580 and 630 nm (580/630 ratio) of SNARF-5F is calculated in real time. As the pH is more acidic, SNARF-5F fluorescence increases at 580 nm while decreases at 630nm. To calibrate the system, Joklik’s EMEM medium titrated at different pH (7.4, 7, 6.5, 6, 5.5 and 5) was added with 4 µM SNARF-5F, emulsified and droplets screened for fluorescence. The 580/630nm ratio were calculated for each pH and plotted against the respective pH, which allowed constructing a calibration curve.

### Cell lines

Breast cancer cell lines MDA-MB-231 and MCF7 were obtained from the American Type Culture Collection and cultured in DMEM medium (Sigma) supplemented with 10% FBS, in a humidified atmosphere at 37 °C and 5% CO2. Cells used were detached before confluency, tested for absence of mycoplasma contamination and validated for short tandem repeat profiling.

### Identification of a pH threshold to discriminate CTC from WBC in patient samples

To identify a threshold of extracellular acidification rate (i.e.: pH) able to better discriminate between CTC and WBC, the ECAR of breast cancer cell lines (MDA-MB-231 and MCF7), and WBCs was separately assessed by using the MBA. WBCs were obtained from the peripheral blood of healthy donors (n = 3), after red blood cell lysis with a 1X FACS lysing solution (BD Pharm Lyse), while breast cancer cell line (MCF7 and MDA-MB-231), grown as previously described at a confluence of 70-80%, were detached with 1% Trypsin-EDTA. Cells were resuspended at a concentration of 1,000,000 cells/ml in 50 μl of the unbuffered Joklik’s modified EMEM culture medium (Sigma) containing 2mM EDTA, 0.1% BSA, 15% Optiprep and 4mM of the fluorescent pH indicator SNARF-5F, emulsified as described in the previous section, incubated at 37°C for 30 min and analyzed. All experiments were performed in triplicate at various cell passages.

### CTC detection by the MBA

Blood samples were drawn into K_2_-EDTA Vacutainer tubes (Beckton Dickinson) and maintained at room temperature. For each sample, 2.5 ml of blood were analyzed within 2 hours after collection. Red blood cells (RBC) were lysed with a 1X FACS lysing solution (BD Pharm Lyse) and centrifuged at 200 RCF for 5 min. Thereafter, the nucleated fraction was depleted of CD45-positive WBC and residual RBC using CD45 and Glycophorin A microbeads (Miltenyi Biotec), respectively, and LD separation columns in a MACS MIDI separator (Miltenyi Biotec), according to the manufacturer’s instructions. The CD45-negative cells were recovered, centrifuged at 300 RCF for 10 min and stained with anti-CD45 (BD Horizon Brilliant™ Violet 480, dilution 1:100) and anti-EpCAM (BD Horizon Brilliant™ Violet 421, dilution 1:100) for 25 min at room temperature. After washing the sample with PBS-BSA 0.5%, cells were resuspended in 50 μl of the unbuffered Joklik’s modified EMEM culture medium (Sigma) containing 2mM EDTA, 0.1% BSA, 15% Optiprep and 4mM of the fluorescent pH indicator SNARF-5F (Thermo Fisher Scientific). Then, cells were single-cell encapsulated in monodispersed droplets using the droplet microfluidic platform as described in previous section. The emulsion was collected and incubated at 37L C and 5% CO2 for 30 min and then reinjected into the microfluidic channel for fluorescent reading of pH, CD45 and EpCAM expression, and the acquisition of bright-field imaging of each positive event. Positive events were defined as CD45-negative cells able to acidify their extracellular environment to a pH lower than 6.4 (MBA-CTC). The number of CTC was then proportionally adjusted to 7.5mL for an easier comparison to the one reported by the CellSearch test. This was done by obtaining the total volume of sample analyzed (total volume = droplet volume * number of droplets analyzed) and doing the proper proportion (MBA-CTC (measured) / volume analyzed = MBA-CTC / 7.5mL). All evaluations were performed without knowledge of the clinical status of the patients.

### CTC detection by CellSearch

Blood samples were drawn into 10 mL CellSave Preservative Tubes (Menarini) and maintained at room temperature. Blood samples were sent to Veneto Institute of Oncology IOV-IRCCS, processed and analyzed according to manufacturer’s instruction *(13)*. To quantify the fraction of apoptotic CTCs, a FITC-conjugated anti-M30 monoclonal antibody was used in conjunction with the standard CTC kit, for recognizing a neoepitope in cytokeratin 18 (CK18) that becomes available at a caspase cleavage event during apoptosis, and is undetectable in viable epithelial cells as previously described *(41)*. All evaluations were performed without knowledge of the clinical status of the patients.

### Statistical analysis

Data were analyzed using GraphPad Prism 6 (version 2.6). Patient and clinical characteristics were presented as frequency and percentage, median and range, mean and standard deviation (SD), as appropriate. Comparison of median between groups were performed by Mann-Whitney test and groups were compared using the Wilcoxon rank test, as appropriate. PFS and OS (time elapsed from enrolment to disease progression and death from any cause, respectively) were determined by Kaplan–Meier plots, with data being censored at last follow-up if progression or death had not occurred. Gehan-Breslow-Wilcoxon test were used to compare the survival curves by CTC detection groups. p < 0.05 was considered significant.

### Cut-off determination for ECAR method

Median PFS times and Cox proportional hazard ratios over all possible CTC cut-offs using baseline data were used to determine the optimal CTC cut-off for the prediction of PFS. The median PFS for the patients above or below each cut-off was calculated. In addition, the percentage of patients above each CTC cut-off was calculated. The best cut-off was selected as the one with the highest Cox hazard ratio, being above the normal background and having at least 40% patients above the cut-off. A CTC count of 6 or more per 7.5mL of blood is predictive of shorter PFS. The chart showing median PFS for each cut-off is shown in fig. S4. The cut-off of 6 CTC/7.5mL was determined using only results from the baseline and PFS as the outcome. Additional analyses have shown that the cut-off for other variable and time points (OS at the baseline, PFS and OS at 1^st^ follow-up) may differ. In order to show uniform results and simplify the interpretation, a CTC cut-off of 6 CTC/7.5mL was used for all analyses.

## Data Availability

All data are available in the text and supplementary materials.

## SUPPLEMENTARY MATERIALS

Fig. S1. CONSORT diagram describing the study design, enrolment of patients and motivations of data exclusion.

Fig. S2. Principle of the metabolic-based assay (MBA) CTC detection. Fig. S3. Measurement of ECAR by the metabolism-based assay (MBA).

Fig. S4. Determination of the prognostic CTC cut-off for the metabolic-based assay (MBA).

Fig. S5. CTC count of healthy donors (HD) and mBC patients at baseline (T0) and follow-up (T1) performed with the metabolic-based assay (MBA).

Fig. S6. The changes of CTC count before and after therapy.

Table S1. Prevalence of CTCs: comparison between the metabolic-based assay (MBA) and the CellSearch (CS).

Table S2. CTC count in 26 healthy donors as detected with the metabolic-based assay (MBA).

Table S3. CTCs and their correlation with therapy response as assessed by imaging.

## Acknowledgements

We are grateful to the staff of CRO-biobank for their support in patient recruitment and sample management/preparation. We are grateful to the physicians and nurses of the Department of Medical Oncology at CRO. We are also grateful to the patients who participated in this project. We are grateful to Dr. Barbara Belletti (CRO Aviano) for insightful comments on the manuscript.

## Author contributions

F.D.B., M.T., A.C., and R.Z., conceptualization; G.B. and E.B. data curation; G.B. and F.D.B. formal analysis; A.C. and A.S. funding acquisition; G.B., E.B., M.B., E.R., C.R., F.D.B. and A.P., investigation; S.M. and F.D.B. managed patient recruitment; G.B., E.B. and F.D.B. methodology; G.B. and F.D.B: Visualization; A.C., R.Z., G.B., E.R., D.C., A.S., F.D.B., M.T., A.P., and W.T.S.H., interpretation of results; G.B. Writing – original draft; all authors, writing-review & editing.

## Data and materials availability

All data associated with this study are present in the paper or the Supplementary Materials.

## REFERENCE AND NOTES

1. J. Ferlay, M. Colombet, I. Soerjomataram, C. Mathers, D. M. Parkin, M. Piñeros, A. Znaor, F. Bray, Estimating the global cancer incidence and mortality in 2018: GLOBOCAN sources and methods., Int. J. Cancer 144, 1941–1953 (2019).

2. G. Siravegna, S. Marsoni, S. Siena, A. Bardelli, Integrating liquid biopsies into the management of cancer., Nat. Rev. Clin. Oncol. 14, 531–548 (2017).

3. C. Alix-Panabières, K. Pantel, Clinical applications of circulating tumor cells and circulating tumor DNA as liquid biopsy., Cancer Discov. 6, 479–491 (2016).

4. E. S. Antonarakis, C. Lu, H. Wang, B. Luber, M. Nakazawa, J. C. Roeser, Y. Chen, T. A. Mohammad, Y. Chen, H. L. Fedor, T. L. Lotan, Q. Zheng, A. M. De Marzo, J. T. Isaacs, W. B. Isaacs, R. Nadal, C. J. Paller, S. R. Denmeade, M. A. Carducci, M. A. Eisenberger, J. Luo, AR-V7 and resistance to enzalutamide and abiraterone in prostate cancer., N. Engl. J. Med. 371, 1028–1038 (2014).

5. H. I. Scher, D. Lu, N. A. Schreiber, J. Louw, R. P. Graf, H. A. Vargas, A. Johnson, A. Jendrisak, R. Bambury, D. Danila, B. McLaughlin, J. Wahl, S. B. Greene, G. Heller, D. Marrinucci, M. Fleisher, R. Dittamore, Association of AR-V7 on Circulating Tumor Cells as a Treatment-Specific Biomarker With Outcomes and Survival in Castration-Resistant Prostate Cancer., JAMA Oncol. 2, 1441–1449 (2016).

6. T. Fehm, V. Müller, B. Aktas, W. Janni, A. Schneeweiss, E. Stickeler, C. Lattrich, C. R. Löhberg, E. Solomayer, B. Rack, S. Riethdorf, C. Klein, C. Schindlbeck, K. Brocker, S. Kasimir-Bauer, D. Wallwiener, K. Pantel, HER2 status of circulating tumor cells in patients with metastatic breast cancer: a prospective, multicenter trial., Breast Cancer Res. Treat. 124, 403–412 (2010).

7. N. Beije, W. Onstenk, J. Kraan, A. M. Sieuwerts, P. Hamberg, L. Y. Dirix, A. Brouwer, F. E. de Jongh, A. Jager, C. M. Seynaeve, N. M. Van, J. A. Foekens, J. W. M. Martens, S. Sleijfer, Prognostic Impact of HER2 and ER Status of Circulating Tumor Cells in Metastatic Breast Cancer Patients with a HER2-Negative Primary Tumor., Neoplasia 18, 647–653 (2016).

8. L. Zhang, L. D. Ridgway, M. D. Wetzel, J. Ngo, W. Yin, D. Kumar, J. C. Goodman, M. D. Groves, D. Marchetti, The identification and characterization of breast cancer CTCs competent for brain metastasis., Sci. Transl. Med. 5, 180ra48 (2013).

9. J. Y. Pierga, D. Hajage, T. Bachelot, S. Delaloge, E. Brain, M. Campone, V. Diéras, E. Rolland, L. Mignot, C. Mathiot, F. C. Bidard, High independent prognostic and predictive value of circulating tumor cells compared with serum tumor markers in a large prospective trial in first-line chemotherapy for metastatic breast cancer patients., Ann. Oncol. 23, 618–624 (2012).

10. F.-C. Bidard, D. J. Peeters, T. Fehm, F. Nolé, R. Gisbert-Criado, D. Mavroudis, S. Grisanti, D. Generali, J. A. Garcia-Saenz, J. Stebbing, C. Caldas, P. Gazzaniga, L. Manso, R. Zamarchi, A. F. de Lascoiti, L. De Mattos-Arruda, M. Ignatiadis, R. Lebofsky, S. J. van Laere, F. Meier-Stiegen, M.-T. Sandri, J. Vidal-Martinez, E. Politaki, F. Consoli, A. Bottini, E. Diaz-Rubio, J. Krell, S.-J. Dawson, C. Raimondi, A. Rutten, W. Janni, E. Munzone, V. Carañana, S. Agelaki, C. Almici, L. Dirix, E.-F. Solomayer, L. Zorzino, H. Johannes, J. S. Reis-Filho, K. Pantel, J.-Y. Pierga, S. Michiels, Clinical validity of circulating tumour cells in patients with metastatic breast cancer: a pooled analysis of individual patient data., Lancet Oncol. 15, 406–414 (2014).

11. G. T. Budd, M. Cristofanilli, M. J. Ellis, A. Stopeck, E. Borden, M. C. Miller, J. Matera, M. Repollet, G. V. Doyle, L. W. M. M. Terstappen, D. F. Hayes, Circulating tumor cells versus imaging--predicting overall survival in metastatic breast cancer., Clin. Cancer Res. 12, 6403–6409 (2006).

12. F.-C. Bidard, T. Fehm, M. Ignatiadis, J. B. Smerage, C. Alix-Panabières, W. Janni, C. Messina, C. Paoletti, V. Müller, D. F. Hayes, M. Piccart, J.-Y. Pierga, Clinical application of circulating tumor cells in breast cancer: overview of the current interventional trials., Cancer Metastasis Rev. 32, 179–188 (2013).

13. S. Riethdorf, H. Fritsche, V. Müller, T. Rau, C. Schindlbeck, B. Rack, W. Janni, C. Coith, K. Beck, F. Jänicke, S. Jackson, T. Gornet, M. Cristofanilli, K. Pantel, Detection of circulating tumor cells in peripheral blood of patients with metastatic breast cancer: a validation study of the CellSearch system., Clin. Cancer Res. 13, 920–928 (2007).

14. L. Zhang, S. Riethdorf, G. Wu, T. Wang, K. Yang, G. Peng, J. Liu, K. Pantel, Meta-analysis of the prognostic value of circulating tumor cells in breast cancer., Clin. Cancer Res. 18, 5701–5710 (2012).

15. M. Cristofanilli, G. T. Budd, M. J. Ellis, A. Stopeck, J. Matera, M. C. Miller, J. M. Reuben, G. V. Doyle, W. J. Allard, L. W. M. M. Terstappen, D. F. Hayes, Circulating tumor cells, disease progression, and survival in metastatic breast cancer., N. Engl. J. Med. 351, 781–791 (2004).

16. D. F. Hayes, M. Cristofanilli, G. T. Budd, M. J. Ellis, A. Stopeck, M. C. Miller, J. Matera, W. J. Allard, G. V. Doyle, L. W. Terstappen, Circulating tumor cells at each follow-up time point during therapy of metastatic breast cancer patients predict progression-free and overall survival., Clin. Cancer Res. 12, 4218–4224 (2006).

17. M. Bulfoni, M. Turetta, F. Del Ben, C. Di Loreto, A. P. Beltrami, D. Cesselli, Dissecting the heterogeneity of circulating tumor cells in metastatic breast cancer: going far beyond the needle in the haystack., Int. J. Mol. Sci. 17 (2016), doi:10.3390/ijms17101775.

18. M. Yu, A. Bardia, B. S. Wittner, S. L. Stott, M. E. Smas, D. T. Ting, S. J. Isakoff, J. C. Ciciliano, M. N. Wells, A. M. Shah, K. F. Concannon, M. C. Donaldson, L. V. Sequist, E. Brachtel, D. Sgroi, J. Baselga, S. Ramaswamy, M. Toner, D. A. Haber, S. Maheswaran, Circulating breast tumor cells exhibit dynamic changes in epithelial and mesenchymal composition., Science 339, 580–584 (2013).

19. A. Satelli, A. Mitra, Z. Brownlee, X. Xia, S. Bellister, M. J. Overman, S. Kopetz, L. M. Ellis, Q. H. Meng, S. Li, Epithelial-mesenchymal transitioned circulating tumor cells capture for detecting tumor progression., Clin. Cancer Res. 21, 899–906 (2015).

20. M. Bulfoni, L. Gerratana, F. Del Ben, S. Marzinotto, M. Sorrentino, M. Turetta, G. Scoles, B. Toffoletto, M. Isola, C. A. Beltrami, C. Di Loreto, A. P. Beltrami, F. Puglisi, D. Cesselli, In patients with metastatic breast cancer the identification of circulating tumor cells in epithelial-to-mesenchymal transition is associated with a poor prognosis., Breast Cancer Res. 18, 30 (2016).

21. M. Poudineh, P. M. Aldridge, S. Ahmed, B. J. Green, L. Kermanshah, V. Nguyen, C. Tu, R. M. Mohamadi, R. K. Nam, A. Hansen, S. S. Sridhar, A. Finelli, N. E. Fleshner, A. M. Joshua, E. H. Sargent, S. O. Kelley, Tracking the dynamics of circulating tumour cell phenotypes using nanoparticle-mediated magnetic ranking., Nat. Nanotechnol. 12, 274–281 (2017).

22. Y. Tang, Z. Wang, Z. Li, J. Kim, Y. Deng, Y. Li, J. R. Heath, W. Wei, S. Lu, Q. Shi, High-throughput screening of rare metabolically active tumor cells in pleural effusion and peripheral blood of lung cancer patients., Proc. Natl. Acad. Sci. USA 114, 2544–2549 (2017).

23. M. Turetta, M. Bulfoni, G. Brisotto, G. Fasola, A. Zanello, E. Biscontin, L. Mariuzzi, A. Steffan, C. Di Loreto, D. Cesselli, F. Del Ben, Assessment of the mutational status of NSCLC using hypermetabolic circulating tumor cells., Cancers (Basel) 10 (2018), doi:10.3390/cancers10080270.

24. J. Chen, S. Cao, B. Situ, J. Zhong, Y. Hu, S. Li, J. Huang, J. Xu, S. Wu, J. Lin, Q. Zhao, Z. Cai, L. Zheng, Q. Wang, Metabolic reprogramming-based characterization of circulating tumor cells in prostate cancer., J Exp Clin Cancer Res 37, 127 (2018).

25. F. Del Ben, M. Turetta, G. Celetti, A. Piruska, M. Bulfoni, D. Cesselli, W. T. S. Huck, G. Scoles, A Method for Detecting Circulating Tumor Cells Based on the Measurement of Single-Cell Metabolism in Droplet-Based Microfluidics., Angew. Chem. Int. Ed. Engl. 55, 8581–8584 (2016).

26. Y. Zhang, Y. Tang, S. Sun, Z. Wang, W. Wu, X. Zhao, D. M. Czajkowsky, Y. Li, J. Tian, L. Xu, W. Wei, Y. Deng, Q. Shi, Single-cell codetection of metabolic activity, intracellular functional proteins, and genetic mutations from rare circulating tumor cells., Anal. Chem. 87, 9761–9768 (2015).

27. H. Cai, F. Peng, 2-NBDG fluorescence imaging of hypermetabolic circulating tumor cells in mouse xenograft model of breast cancer., J Fluoresc 23, 213–220 (2013).

28. D. Hanahan, R. A. Weinberg, Hallmarks of cancer: the next generation., Cell 144, 646–674 (2011).

29. U. E. Martinez-Outschoorn, M. Peiris-Pagés, R. G. Pestell, F. Sotgia, M. P. Lisanti, Cancer metabolism: a therapeutic perspective., Nat. Rev. Clin. Oncol. 14, 11–31 (2017).

30. S. Granja, D. Tavares-Valente, O. Queirós, F. Baltazar, Value of pH regulators in the diagnosis, prognosis and treatment of cancer., Semin. Cancer Biol. 43, 17–34 (2017).

31. S. R. Pillai, M. Damaghi, Y. Marunaka, E. P. Spugnini, S. Fais, R. J. Gillies, Causes, consequences, and therapy of tumors acidosis., Cancer Metastasis Rev. (2019), doi:10.1007/s10555-019-09792-7.

32. G. J. Kelloff, J. M. Hoffman, B. Johnson, H. I. Scher, B. A. Siegel, E. Y. Cheng, B. D. Cheson, J. O’shaughnessy, K. Z. Guyton, D. A. Mankoff, L. Shankar, S. M. Larson, C. C. Sigman, R. L. Schilsky, D. C. Sullivan, Progress and promise of FDG-PET imaging for cancer patient management and oncologic drug development., Clin. Cancer Res. 11, 2785–2808 (2005).

33. A. Mongersun, I. Smeenk, G. Pratx, P. Asuri, P. Abbyad, Droplet Microfluidic Platform for the Determination of Single-Cell Lactate Release., Anal. Chem. 88, 3257–3263 (2016).

34. M. E. Gallina, T. J. Kim, M. Shelor, J. Vasquez, A. Mongersun, M. Kim, S. K. Y. Tang, P. Abbyad, G. Pratx, Toward a Droplet-Based Single-Cell Radiometric Assay., Anal. Chem. 89, 6472–6481 (2017).

35. R. A. Cardone, V. Casavola, S. J. Reshkin, The role of disturbed pH dynamics and the Na+/H+ exchanger in metastasis., Nat. Rev. Cancer 5, 786–795 (2005).

36. B. A. Webb, M. Chimenti, M. P. Jacobson, D. L. Barber, Dysregulated pH: a perfect storm for cancer progression., Nat. Rev. Cancer 11, 671–677 (2011).

37. R. A. Gatenby, R. J. Gillies, A microenvironmental model of carcinogenesis., Nat. Rev. Cancer 8, 56–61 (2008).

38. L. Mazutis, J. Gilbert, W. L. Ung, D. A. Weitz, A. D. Griffiths, J. A. Heyman, Single-cell analysis and sorting using droplet-based microfluidics., Nat. Protoc. 8, 870–891 (2013).

39. M. He, J. S. Edgar, G. D. M. Jeffries, R. M. Lorenz, J. P. Shelby, D. T. Chiu, Selective encapsulation of single cells and subcellular organelles into picoliter- and femtoliter-volume droplets., Anal. Chem. 77, 1539–1544 (2005).

40. A. Martowicz, G. Spizzo, G. Gastl, G. Untergasser, Phenotype-dependent effects of EpCAM expression on growth and invasion of human breast cancer cell lines., BMC Cancer 12, 501 (2012).

41. E. Rossi, U. Basso, R. Celadin, F. Zilio, S. Pucciarelli, M. Aieta, C. Barile, T. Sava, G. Bonciarelli, S. Tumolo, C. Ghiotto, C. Magro, A. Jirillo, S. Indraccolo, A. Amadori, R. Zamarchi, M30 neoepitope expression in epithelial cancer: quantification of apoptosis in circulating tumor cells by CellSearch analysis., Clin. Cancer Res. 16, 5233–5243 (2010).

42. J. S. de Bono, H. I. Scher, R. B. Montgomery, C. Parker, M. C. Miller, H. Tissing, G. V. Doyle, L. W. W. M. Terstappen, K. J. Pienta, D. Raghavan, Circulating tumor cells predict survival benefit from treatment in metastatic castration-resistant prostate cancer., Clin. Cancer Res. 14, 6302–6309 (2008).

43. S. J. Cohen, C. J. A. Punt, N. Iannotti, B. H. Saidman, K. D. Sabbath, N. Y. Gabrail, J. Picus, M. Morse, E. Mitchell, M. C. Miller, G. V. Doyle, H. Tissing, L. W. M. M. Terstappen, N. J. Meropol, Relationship of circulating tumor cells to tumor response, progression-free survival, and overall survival in patients with metastatic colorectal cancer., J. Clin. Oncol. 26, 3213–3221 (2008).

44. S. Taylor, E. P. Spugnini, Y. G. Assaraf, T. Azzarito, C. Rauch, S. Fais, Microenvironment acidity as a major determinant of tumor chemoresistance: Proton pump inhibitors (PPIs) as a novel therapeutic approach., Drug Resist Updat 23, 69–78 (2015).

45. D. Neri, C. T. Supuran, Interfering with pH regulation in tumours as a therapeutic strategy., Nat. Rev. Drug Discov. 10, 767–777 (2011).

